# Salivary bacterial signatures in depression-obesity comorbidity are associated with neurotransmitters and neuroactive dipeptides

**DOI:** 10.1101/2021.08.10.21255754

**Authors:** Gajender Aleti, Jordan N. Kohn, Emily A. Troyer, Kelly Weldon, Shi Huang, Anupriya Tripathi, Pieter C. Dorrestein, Austin D. Swafford, Rob Knight, Suzi Hong

## Abstract

**Background:** Depression and obesity, both of which are highly prevalent and inflammation underlies, often co-occur. Microbiome perturbations are implicated in obesity-inflammation-depression interrelationships, but how microbiome alterations contribute to underlying pathologic processes remains unclear. Metabolomic investigations to uncover microbial neuroactive metabolites may offer mechanistic insights into host-microbe interactions.

**Methods:** Using 16S sequencing and untargeted mass spectrometry of saliva, and blood monocyte inflammation regulation assays, we determined key microbes, metabolites and host inflammation in association with depressive symptomatology, obesity, and depressive symptomatology-obesity comorbidity.

**Results:** Gram-negative bacteria with inflammation potential were enriched relative to Gram-positive bacteria in comorbid obesity-depression, supporting the inflammation-oral microbiome link in obesity-depression interrelationships. Oral microbiome was highly predictive of depressive symptomatology-obesity co-occurrences than obesity and depressive symptomatology independently, suggesting specific microbial signatures associated with obesity-depression co-occurrences. Mass spectrometry analysis revealed significant changes in levels of signaling molecules of microbiota, microbial or dietary derived signaling peptides and aromatic amino acids among host phenotypes. Furthermore, integration of the microbiome and metabolomics data revealed that key oral microbes, many previously shown to have neuroactive potential, co-occurred with potential neuropeptides and biosynthetic precursors of the neurotransmitters dopamine, epinephrine and serotonin.

**Conclusions:** Together, our findings offer novel insights into oral microbial-brain connection and potential neuroactive metabolites involved.

## Background

Depression and obesity are common, debilitating, and frequently co-occurring chronic conditions with increasing incidences globally [1]. Nearly 39% of the adult population are overweight and 13% are obese worldwide (WHO, 2016), while 5% of the world population are affected by mood disorders (WHO, 2017) [2, 3]. The relationship between obesity and depression is often bidirectional [4], as prevalence of depression among individuals with obesity is significantly higher than that in the general population [5, 6]. Conversely, individuals with depression are more likely to develop obesity compared to non-depressed individuals [7]. Despite the advent of antidepressant drugs and their long-term usage in clinical treatment, the majority of patients with depression are treatment-refractory, and obesity may further reduce the efficacy of antidepressants [8]. Furthermore, comorbid depression and obesity are strongly associated with several diseases such as type 2 diabetes mellitus, cardiovascular diseases, chronic kidney disease and cancer, reducing both longevity and quality of life [2, 9]. Therefore, obesity and depression, and their co-occurrence, pose a major public health concern worldwide.

Inflammatory dysregulation is a common pathogenic mechanism underlying the co- occurrence of depression and obesity, as both are associated with chronic low-grade inflammation [10, 11]. Individuals with obesity and depression evidence increased concentrations of peripheral and central inflammatory cytokines and acute phase reactants, such as interleukin (IL)-6, tumor necrosis factor alpha (TNF-α), and C-reactive protein (CRP) [11, 12]. In obesity, macrophages accumulate in adipose tissue leading to local and systemic inflammation [13, 14], which can contribute to depressive symptoms via multiple mechanisms, such as by decreasing neurotransmitter availability, and by potentiating neuroinflammatory processes such as microglial activation and peripheral monocyte trafficking to the central nervous system (CNS) [10,15,16]. It should be noted, however, that inflammation has been shown to underlie only a subset of depression cases [17], hence the conceptualization of a theoretical immuno-metabolic *subtype* of major depressive disorder [18]. Nonetheless, inflammatory dysregulation remains a central mechanism underlying the co-occurrence of depression and obesity, and this is likely relevant to sub-clinical depressive symptomatology. To this end, our previous work has demonstrated that even in individuals without clinical diagnosis of depression, higher depressive symptom scores, obesity, and downregulated glucocorticoid and adrenergic receptor-mediated cellular inflammatory control are interrelated [19–21].

Although psychological stress, host genetics and environmental factors have been shown to contribute to obesity and depression, recently, the human microbiome (i.e., collection of diverse microorganisms and their genetic material) and metabolome (i.e., a large collection of structurally diverse metabolites) have been implicated in processes of energy homeostasis, mood and behavior, and immune regulation, and may therefore offer a novel mechanism underlying the co-occurrence of depression and obesity [2]. Animal studies of obesity have shown that depletion of members of *Bifidobacterium*, *Lactobacillus*, and *Akkermansia* are associated with weight gain, increased inflammation, increased depressive behavior and changes in neural circuitry [22, 23]. Animal studies have also shown that increased permeability in the intestinal barrier and the blood-brain barrier (BBB) are associated with increased plasma lipopolysaccharide (LPS) levels [22–24] and neuroinflammation [23]. Altogether, these studies suggest that increased intestinal barrier permeability and subsequent translocation of gut bacterial endotoxin, particularly LPS from Gram-negative bacterial cell walls, into systemic circulation, is a source of chronic low-grade inflammation and metabolic endotoxemia, which can potentiate neuroinflammatory processes, and therefore serve as a potential mechanism underlying the occurrence of depressive symptoms in the context of obesity. However, this remains to be established in humans.

It is to be noted that human microbiome studies in depression and obesity, and indeed in health and disease, have focused largely on the ecosystem of the distal gut, while few studies have examined the microbial ecology of the oral cavity outside of oral-related conditions such as dental caries (i.e., tooth decay) and periodontitis (i.e., severe gum inflammation). The oral cavity, an entry portal to both the digestive and respiratory tracts, contains the most diverse microbial community after the gut, harboring more than 700 unique bacterial species with at least 150 specialized bacterial species per mouth [25, 26]. More than 60% of the microbial species found in the oral cavity have been shown to be potentially transmitted to the gut, suggesting that oral cavity is a reservoir for gut microbial strains in shaping the gut microbiome in health and disease [27]. Dysregulation of the unique microbe-microbe and microbe-host interactions in the oral ecosystem has been associated with systemic inflammatory diseases such as inflammatory bowel syndrome [28, 29] beyond an array of oral diseases. In addition, oral microbiota have also been associated with several neurological diseases, such as Alzheimer’s disease (AD) [30], multiple sclerosis [31] and Parkinson’s disease [32]. Previously, our group found that salivary microbial diversity and diurnal variability were associated with both peripheral proinflammatory cytokine levels and psychological distress in this cohort on which this study is based [33]. The intimate link between the oral microbiota and systemic human diseases, as evidenced by aforementioned studies suggests that the oral cavity is likely a promising site for gaining insight into the pathophysiology of depression-obesity comorbidity. Moreover, the oral cavity is easily accessible via non-invasive as well as ‘on-demand’ collection of saliva samples for multi-omics applications.

While mechanisms linking the oral microbiota to the brain (i.e. “oral-brain axis”) remain largely unknown [34, 35], recent studies have speculated several transmission routes of how oral bacteria may reach the brain and influence neuro-immune activity and inflammation [36]. For instance, routine dental procedures such as flossing, brushing and cleaning may cause oral bacteria to enter the blood circulation and cause bacteremia [37], and some of these microbes may traverse the BBB. Alteration in the permeability of the BBB may also expose the brain to bacterial metabolites triggering an inflammatory response, which in turn alters functioning of the CNS. For example, *Porphyromonas gingivalis*, a resident oral bacterium and a keystone pathogen in periodontitis has been found in the brain of AD patients [30] as well as neurotoxic proteases i.e., gingipains produced by *P. gingivalis* [30].

A recent study has shown that human gut bacteria encode at least 56 gut-brain metabolic pathways, which encompass both known and novel microbial pathways for synthesis and degradation of a number of neurotransmitters that have potential to cross the intestinal barrier and BBB [35]. A subset of these gut-brain pathway effectors, for instance dopamine, glutamate, tryptophan and gamma-aminobutyric acid (GABA) were either enriched or depleted in patients with major depression [35]. In particular, tryptophan metabolic pathways have been shown to be widely distributed across human gut bacterial species [35]. Intriguingly, the majority of these gut bacterial species with neuroactive potential are also found to be residents of the oral cavity [25]. However, to what extent these bacterial species can truly biosynthesize neurotransmitters within the host, either in the gut or the oral cavity, remains unknown. Thus, utilization of metabolomics offers a functional readout of both host and microbial phenotypes encoded in the genome [38, 39], and in conjunction with microbiome analyses, can provide mechanistic insights, yet current knowledge is greatly limited. In particular, microbial specialized metabolites have been shown to be canonical mediators of microbe-microbe and microbe-host interactions, and the most predominant specialized metabolites are of great interest for understanding the mechanisms of these interactions at the molecular level [38–40]. In this regard, the vast and highly diverse array of short peptides shown to play key roles in bacterial cell signaling [41], immune modulation, and neuroactive metabolism [42–44] remains largely unexplored. A recent study has shown that depletion of a variety of structurally uncharacterized dipeptides are associated with inflammatory bowel disease, a chronic inflammatory condition of the gastrointestinal tract [45]. These observations prompted us to hypothesize that neurotransmitters and dipeptides likely have pivotal roles in obesity-inflammation-depression interrelationships.

In this study we aimed to investigate whether oral microbiota and small-molecule mediators of key microbe-microbe and microbe-host interactions differ by depressive symptomatology and obesity as well as their co-occurrence, and are influenced by inflammatory processes. We performed 16S rRNA gene-based sequencing of the oral microbiome and untargeted mass spectrometry of small-molecules from saliva, as well as host inflammation regulation profiles in blood from 60 participants.

## Methods

### Participants

A total of 60 lean to obese participants (20-65 years old) with a range of subclinical depressive symptoms, participating in a larger study investigating the impact of obesity on vascular inflammation and immune cell activation in normotension versus stage 1 hypertension (Basal systolic blood pressure (BP): 130-140 mmHg and diastolic BP: 80-90 mmHg), were included in this study and provided saliva samples. Participant inclusion/exclusion criteria were previously described in detail [33]. Briefly, participants were excluded if they had diabetes, recent history of smoking or substance abuse, history of cardiovascular disease, history of bronchospastic pulmonary disease, inflammatory disorders or health-related factors affecting immune function, psychosis, major depressive disorder, and stage 2 clinical hypertension or with average BP ≥145/90 mmHg measured at the lab visit from six measurements on two separate days, using a Dinamap Compact BP monitor (Critikon, Tampa, FL). Sociodemographic characteristics (i.e., age, sex, and race) and anthropometrics (i.e., height, weight, hip and waist circumference) data were collected.

### Obesity characterization

BMI was calculated based on height and weight measurements (kg/m^2^), and individuals were dichotomized into two groups, based on our prior findings of little notable differences in inflammatory or depressive symptoms state between lean and overweight individuals (ref): non- obese (BMI <30 kg/m^2^) and obese (BMI ≥30 kg/m^2^). For further adiposity characterization dual x-ray absorptiometry was performed to calculate %total and trunk body fat.

### Depressive symptomatology assessment

Depressive symptoms were assessed using the Beck Depression Inventory (BDI-Ia), a comprehensive and clinically robust self-report 21-item questionnaire (Beck et al., 1996). Each question was scored from 0-3, summed to a BDI total score (BDI-T), and then subcategorized into cognitive-affective (BDI-C) and somatic (BDI-S) depression scores based on the items such as BDI-C: guilt, pessimism and BDI-S: fatigue, sleep disruption [46].

Based on obesity status and mean BDI-T scores, participants were categorized into 4- groups: non-obese and lower-depressive controls (N=10 participants; n=43 samples; “controls”), obese and lower-depressive (N=18; n=74; “Ob/lower-Dep”), non-obese and higher-depressive symptoms (N=5; n=22; “Non-ob/higher-Dep”), and obese and higher-depressive symptoms (N=27; n=122; “Ob/higher-Dep”).

### Blood collection and cellular inflammation assay

For detailed protocol, see Supplementary Materials and Methods section. Briefly, LPS- stimulated blood was incubated with beta-adrenergic receptor agonist isoproterenol and evaluated for intracellular monocyte TNF-α production using flow cytometry, as previously described [47]. Monocyte beta-adrenergic receptor-mediated inflammation control (i.e., “BARIC”, a measure of systemic inflammation) was calculated as the arithmetic difference in %TNF-α-producing monocytes between LPS + media-treated and LPS + isoproterenol-treated samples.

### Saliva collection, DNA extraction and 16S sequencing

For detailed protocols of saliva collection procedure and 16S analysis, see Supplementary Materials and Methods section. Saliva from each participant was collected at five time points across a single day: waking, mid-morning (10:00 hrs), midday (12:00 hrs), afternoon (14:00 hrs), and evening (17:00 hr).

### Statistical analyses

Statistical analyses were conducted using R software (version 3.6.3) in RStudio (version 1.2.5019). First, associations among continuous and categorical metadata variables i.e., age, obesity (BMI, %total body fat and trunk fat), BARIC, BDI scores (BDI-T, BDI-C and BDI-S) were assessed using univariate Spearman correlations across all participants using *psych* package in R software. We applied a simple linear mixed-effects model (LMM) fit to model two alpha diversity measures (Shannon index and Faith’s PD) using restricted maximum likelihood (REML) with a random intercept by participant to account for repeated measurements across the day, and main effects of obesity status, depressive symptom status, and BARIC. Age, sex, race were included as covariates in the model. Beta-diversity between groups was tested using non- parametric *PERMANOVA* with 999 permutations constrained by participant to adjust for 3-5 samples per participant, and a test of homogeneity of dispersion was conducted with the same constraints using *PERMDISP2* in *vegan* package to test overall species composition differences within the groups. Next, post-hoc pairwise comparison was performed using *pairwiseAdonis*.

### Random forest classifications

A random forest sample classifier was trained based on the 16S data with tuned hyperparameters (num.trees=500, mtry=45) in the 20-time repeated, stratified 5-fold cross-validation using *caret* package in R software. The dataset was repeatedly split into five groups with similar class distributions, and we trained the classifier on 80% of the data, and made predictions on the remaining 20% of the data in each fold iteration. We next evaluated the performance of the classifier on predicting the four groups (i.e. controls, Ob/lower-Dep, Non-ob/higher-Dep, Ob/higher-Dep) using both area under the receiver operating characteristic curve (AUROC) and area under the precision-recall curve (AUPRC) based on the samples’ predictions in the holdout test set using *PRROC* package in R. To account for multiple samples per-participant, we next performed 20-time repeated group 3-fold cross-validation, where each participant is in a different testing fold and also samples from the same subjects are never in both testing and training folds.

### Small molecule metabolite detection through mass spectrometry

Saliva was dried and resuspended in 80% MeOH−20% water and submitted to untargeted LC/MS/MS analysis. For a detailed protocol, see Supplementary Materials and Methods section. To examine the metabolic potential in the oral ecosystem and understand the intimate link between salivary microbiota and metabolome in obesity-depressive symptom relationships, we conducted untargeted liquid chromatography-tandem mass spectrometry (LC-MS/MS) analysis of the saliva samples from the same participants who were first investigated for taxonomic profiling in the above analyses [48, 49]. By integrating feature based molecular networking [50] with an automated chemical classification [51] and reference frame based differential abundance analysis [52] approaches, we revealed differential representation of the key molecular features in obesity and depressive symptom conditions.

### Feature based mass spectral molecular networking (FBMN) and chemically-informed comparison of metabolomic profiles

A data matrix of MS1 features that triggered MS2 scans were uploaded along with the metadata file to Global Natural Product Social Molecular Networking (GNPS) (https://gnps.ucsd.edu) [49]. Feature-based molecular networking (version release_20) [50] was performed, and library IDs were generated (see Supplementary Materials and Methods section). To further gain a broad overview of the chemistry of salivary metabolomes from MS/MS data, utilizing an automated chemical classification approach [51], available via GNPS platform, we performed a chemically-informed comparison of untargeted metabolomic profiles across the four groups.

### Differential ranking of taxa and metabolomic features

Differential ranks of taxa and metabolomic features were calculated using Songbird [52], which uses reference frames. Age, sex, race and time of day of saliva collection were provided as covariates in generating a multinomial regression model based on microbial features. Differential microbial features were visualized alongside *de novo* phylogenetic tree constructed from the representative sequences of amplicon sequence variants (ASVs) obtained in this study using EMPress [23]. Statistical significance was tested by applying LMMs on log-ratios of the top-and bottom-20 ranked microbes for each group obtained using Qurro rank plots [53]. We applied a linear regression model by utilizing log-ratios of bacterial features and BARIC inflammatory scores to test interactions between obesity-depressive symptoms and inflammation relationships.

To mitigate the inter-batch effect often observed in the metabolomics data due to technical limitations in the number of samples processed in a batch, relative abundances were adjusted for batch specific-effect along with age, sex, race and time of day, utilizing the multivariate model in the reference frame-based approach [52]. We chose cluster 1 (90 features) as the denominator (“reference frame”) for the log-ratio calculations due to its high prevalence across samples, and moreover, GNPS analyses groups structurally similar molecules into a cluster. Statistical significance was tested by applying Friedman test to account for repeated measurements, prior to multiple pairwise comparison analysis using Wilcoxon rank-sum tests.

### Microbe-metabolite interactions through their co-occurrence probabilities

Permutation based differential abundance testing was performed using discrete false-discovery rate correction method [54] in Calour (https://github.com/biocore/calour) to remove batch-specific MS1 molecular features. Annotated features that were not identified as batch-specific were included in the co-occurrence analysis. Using ASV (N=1516) and annotated molecular features (N=155) as inputs to train neural networks [55] in QIIME 2 [56], we estimated the conditional probability that each molecule is present given the presence of a specific microorganism. The resulting conditional probability matrix representing microbe-metabolite interactions was visualized as an EMPeror biplot [55].

## Results

### Participant characteristics

A total of 261 saliva samples collected from five time points across the day from 60 participants were analyzed (20 – 65 years): 50 participants had five; 51 had four, and 54 had three samples which were adjusted in analyses (See Statistical Analyses). Participants were categorized into the following four groups: non-obese (BMI <30 kg/m2) and lower-depressive controls (N=10 participants; n=43 saliva samples; “controls”), obese (BMI ≥30 kg/m2) and lower-depressive (N=18; n=74; “Ob/lower-Dep”), non-obese and higher-depressive symptoms (N=5; n=22; “Non-ob/higher-Dep”), and obese and higher-depressive symptoms (N=27; n=122; “Ob/higher-Dep”). Sociodemographic characteristics are presented across participant groups (Table 1).

**Table 1.** Demographic and clinical characteristics of participants.

| Variable | Non-obese low depressive<sup>a</sup> | Obese low depressive<sup>b</sup> | Non-obese high depressive<sup>c</sup> | Obese high depressive<sup>d</sup> |
| --- | --- | --- | --- | --- |
| Age | 39±12.2 | 38.9±17.2 | 42.7±10.5 | 43.5±10.9 |
| Sex (%female) | 44 | 50 | 61.1 | 73.3 |
| Race(%C/AA/Asn/NS) | 72/16/12/0 | 37.5/37.5/12.5/12.5 | 55.6/16.7/27.8/0 | 46.7/40/13.3/0 |
| BARIC | 32.1±10.2<sup>d</sup> | 21.9±6.2<sup>c</sup> | 31.8±9<sup>cd</sup> | 25.3±7.5<sup>ac</sup> |
| BMI (kg/m<sup>2</sup>) | 25.1±2.9<sup>bd</sup> | 35.5±4.7<sup>ac</sup> | 26.6±2.9<sup>bd</sup> | 36±4.7<sup>ac</sup> |
| BDI-T | 0.5±0.8<sup>cd</sup> | 0.6±0.7<sup>cd</sup> | 7.9±5.4<sup>ab</sup> | 7.9±5<sup>ab</sup> |

Values presented as mean ± SD. Significant differences between groups were evaluated by Mann-Whitney test and presented as superscripts. Abbreviations: C = Caucasian; AA = African- American; Asn = Asian; NS = Mixed or not specified; BARIC = monocyte beta-adrenergic receptor-mediated inflammation control; BMI = body mass index; BDI-T = Beck Depression Inventory (BDI-Ia) total score.

### Obesity is associated with depressive symptomatology and inflammation

Given that individuals with a clinical diagnosis of depression and/or use of antidepressants were excluded from the study to focus on inflammation-related subclinical depressive symptoms in relation to obesity among otherwise healthy adults, BDI total scores (BDI-T) on average were low (median=3; sd=5; range=0-22). The median value of BDI-T of ≥3 was used to divide participants with relatively ‘higher’ or ‘lower’ depressive symptoms in this non-clinical sample.

In all individuals, BMI was positively correlated with BDI-T scores (*r*=0.29, *p*=0.04), as well as cognitive-affective (*r*=0.27, *p*=0.03) and somatic symptom scores with small to medium effects (*r*=0.22, *p*=0.08) (Figure S1). BARIC values, an indicator of neuro-inflammation regulation, were negatively correlated with BMI (*r*=-0.38, *p*=0.009), and an estimation of adipose tissue volume indicated by %trunk fat (*r*=-0.25, *p*=0.034) across all participants (Figure S1). Age did not moderate any of these relationships, which is in agreement with previous findings [20]. Altogether, obesity was significantly associated with both inflammation regulation and depressive symptoms. However, no significant associations were observed between BARIC and BDI scores in this study (Figure S1).

### Oral microbiota differ based on obesity-depressive symptom groups and inflammation status

Principal coordinates analysis (PCoA) and post-hoc pairwise comparisons of unweighted-UniFrac distances of samples revealed that oral microbiota composition was distinct by obesity (PERMANOVA pseudo-F=0.004, *p*=0.001, Figure 1A, Table 2), BDI-T (PERMANOVA pseudo-F=0.001, *p*=0.0, Figure 1B, Table 2) and across the four obesity-depressive symptom comorbid groups (i.e, Ctrl, Ob/lower-Dep, Non-ob/higher-Dep, Ob/higher-Dep) (Figure 1C, Table 2 and Table 3). Beta-diversity was also significantly differentiated based on the host inflammation across all participants (PERMANOVA pseudo-F=4.71, *p*<0.001, Figure 1D and Table 2). Significant beta-diversity differences were also observed by age, sex, and race but not by sampling time of day (Table 2). Phylogenetic alpha-diversity increased with inflammation (Faith’s PD: t=-2.312, *p*=0.025). Inflammation had slightly larger effects (R^2^=0.02) on microbiome composition than obesity (R^2^=0.008) and depressive symptomatology (R^2^=0.01) (Table 2).

**Figure 1.**
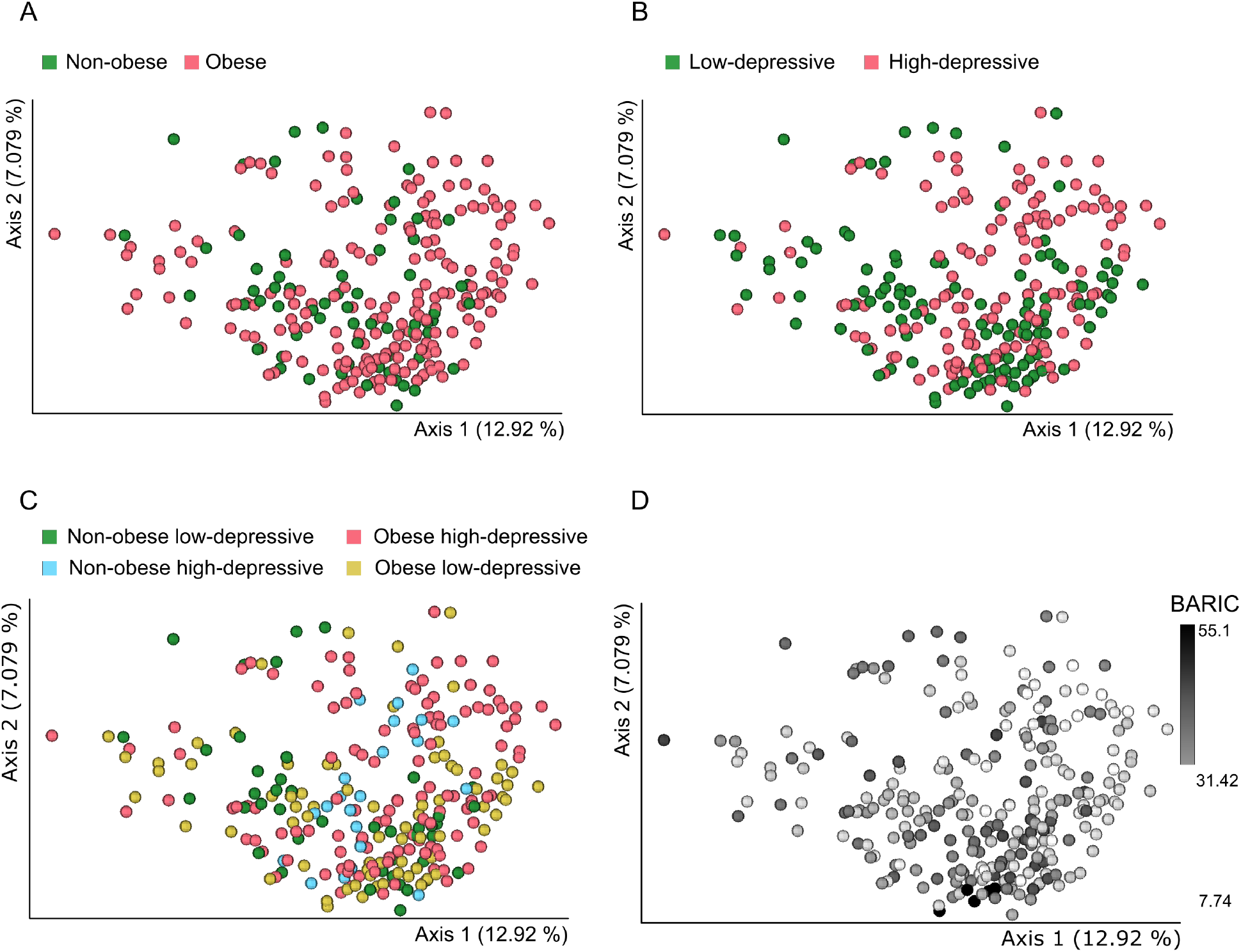
Principal coordinates analyses (PCoA) of oral bacterial communities in (A) non-obese and obese (B) low depressive and higher depressive (C) non-obese low-depressive, non-obese high-depressive, obese, and co-occurring obesity and depressive symptom groups, and (D) in inflammation status. Unweighted-UniFrac distances among samples were visualized using EMPeror. Significance of separation between the groups and further post-hoc pairwise comparisons between groups was tested by applying PERMANOVA test on the principal coordinates.

**Table 2.** Beta-diversity analysis of 16S derived ASVs across groups.

| Variable | Unweighted-UniFrac R<sup>2</sup> | Unweighted-UniFrac F |
| --- | --- | --- |
| Age | 0.01 | 3.88*** |
| Sex | 0.01 | 2.54*** |
| Race | 0.03 | 2.92*** |
| Time of day | 0.01 | 0.98 |
| BARIC | 0.02 | 4.71*** |
| Obesity | 0.008 | 0.004** |
| Depressive symptomatology | 0.01 | 0.001*** |
| Obesity-depressive symptomatology co-occurrences | 0.03 | 2.48*** |

Asteriks indicate statistical significance of PERMANOVA test, p<0.05.

**Table 3.**
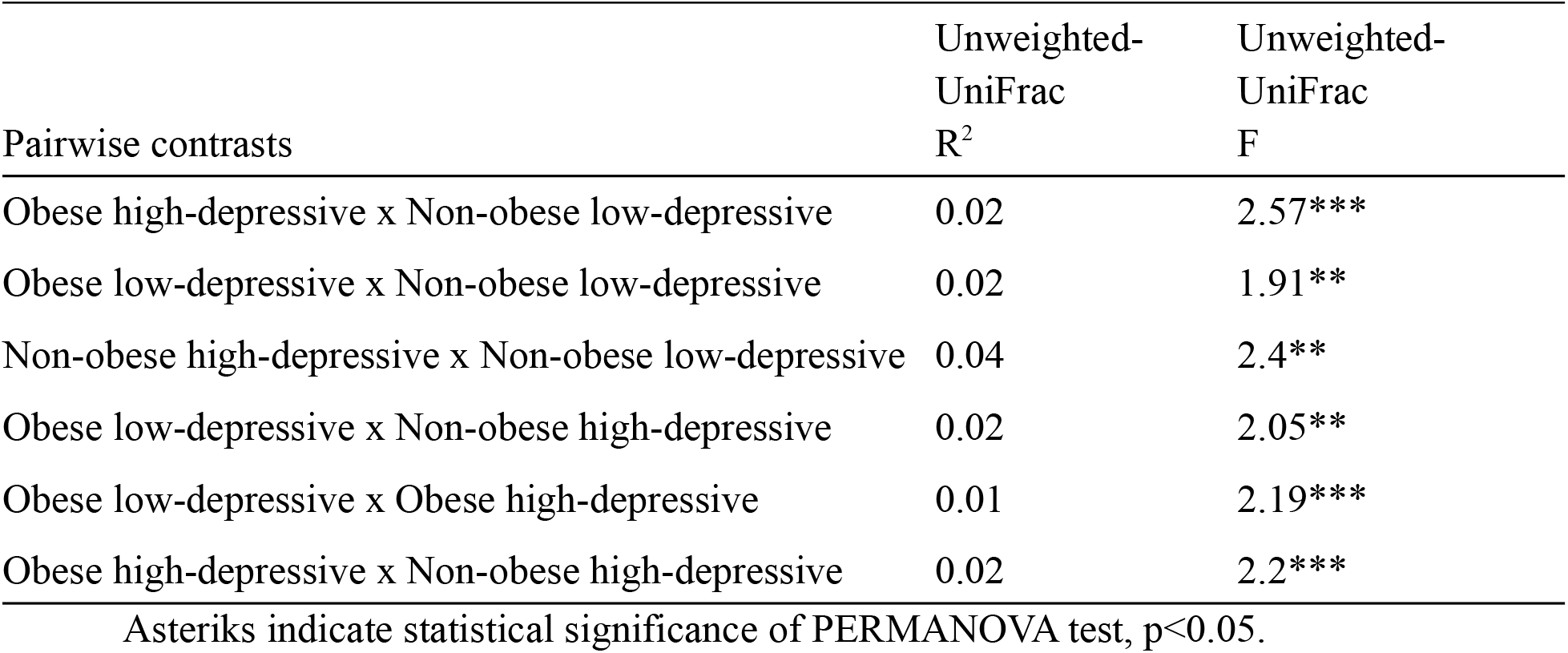
Post-hoc pairwise comparisons of beta-diversity between groups.

| Pairwise contrasts | Unweighted-UniFrac<br>R<sup>2</sup> | Unweighted-UniFrac<br>F |
| --- | --- | --- |
| Obese high-depressive x Non-obese low-depressive | 0.02 | 2.57*** |
| Obese low-depressive x Non-obese low-depressive | 0.02 | 1.91** |
| Non-obese high-depressive x Non-obese low-depressive | 0.04 | 2.4** |
| Obese low-depressive x Non-obese high-depressive | 0.02 | 2.05** |
| Obese low-depressive x Obese high-depressive | 0.01 | 2.19*** |
| Obese high-depressive x Non-obese high-depressive | 0.02 | 2.2*** |

802 Asteriks indicate statistical significance of PERMANOVA test, p<0.05.

### Oral microbiota is predictive of the host obesity-depressive symptomatology

To assess the predictive capacity of the oral microbiome in stratifying individuals with depressive symptoms, obesity and depressive symptomatology-obesity co-occurrence status, we utilized supervised random forest classification. The prediction performance of the model indicated by both area under the receiver operating characteristic curve (AUROC) and area under precision recall curve (AUPRC), revealed high prediction accuracy (AUROC=0.75 and AUPRC=0.74) for obesity-depressive symptom status (Ob/higher Dep) than other groups when multiple samples per-participant were taken into account (Figure 2A and 2B). The Ctrl group was predicted with AUROC=0.75 and AUPRC=0.58; Ob/lower Dep status with AUROC=0.70 and AUPRC=0.49; Non-ob/higher Dep with AUROC=0.70 and AUPRC=0.46. However, at sample-level both AUROC and AUPRC ranged from 0.93 to 0.97, across all groups (Figure S2A and S2B). Altogether, oral microbiome was highly predictive of depressive symptomatology- obesity co-occurrences than obesity and depressive symptomatology independently.

**Figure 2.**
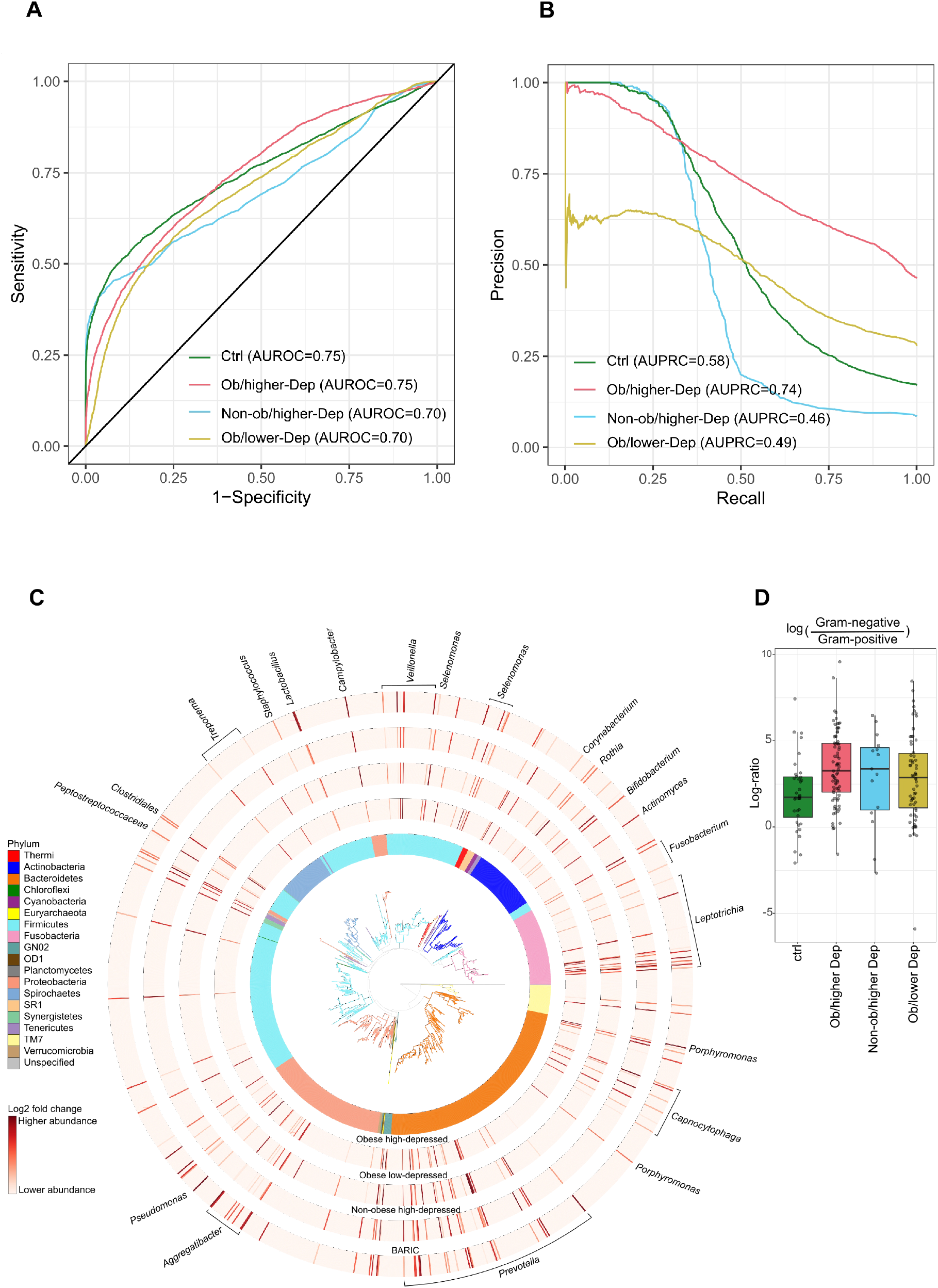
Oral microbiota is distinctly impacted by the host status in co-occurring obesity- depressive status. (A) Receiver operating characteristic curves (AUROC) illustrating classification accuracy of the random forest model across all groups (i.e. controls, Ob/lower Dep, Non-ob/higher-Dep, Ob/higher-Dep). (B) Area under precision recall curves (AUPRC) illustrating performance of the random forest model across all groups. (C) Phylogenetic distribution of the most differentially ranked taxa across the groups. Branches of the *de novo* phylogenetic tree and the innermost ring are colored by phyla. Each barplot layer represents log- fold change abundances of taxa within the group in comparison to the healthy controls i.e. Non- ob/lower-Dep. A multinomial regression model was employed for regressing log-fold change abundances against BARIC values. (D) Log-fold change abundances of Gram-negative microbes relative to Gram-positive microbes across host phenotypes.

### Key oral bacterial taxa are associated with specific host phenotype

Next, we identified the most differentially ranked microbes (99 unique taxa) associated with host phenotypes (Figure 2C). Linear mixed-effects model revealed significant differences in the relative abundances of microbes associated with Ob/higher-Dep (t=6.5, *p*=5.07e-08), Non-ob/higher-Dep (t=-4.2, *p*=0.0002) and Ob/lower Dep (t=-4.5, *p*=5.07e-05) in comparison to Ctrl group, and with inflammation status (t=-4.83, *p*=3.03e-05). Most differentially represented taxa (84 unique taxa) were assigned to Gram-negative bacteria such as *Prevotella*, *Aggregatibacter*, *Pseudomonas*, *Campylobacter,* Clostridia *(Selenomonas, Butyrivibrio*, *Veillonella, Megasphaera* and *Schwartzia*), *Leptotrichia, Capnocytophaga*, and periodontal pathogens such as *Treponema*, *Veillonella*, *Porphyromonas* and *Fusobacterium*. Gram-positive (15 unique taxa) were assigned to Peptostreptococcaceae, Clostridia (*Catonella*, Mogibacteriaceae), *Staphylococcus*, *Corynebacterium*, *Rothia*, *Actinomyces*, and beneficial/probiotic genera *Bifidobacterium* and *Lactobacillus* (Figure 2C, log-fold change abundances for each microbe are shown in Table S1). The Ob/higher-Dep group exhibited a slightly higher abundance of Gram-negative bacteria relative to Gram-positive compared to the Ctrl group (Wilcoxon test: *p*=0.004) (Figure 2D), which were not significantly associated with BARIC scores (data not shown).

### Small molecules detected in saliva are associated with obesity-depressive symptom- inflammation relationships

Untargeted LC-MS/MS analysis of the saliva samples was performed to examine the metabolic potential in the oral ecosystem and understand the intimate link between salivary microbiota and metabolome in obesity-depressive symptom relationships.

The most predominant chemical classes identified from automated chemical classification [51] of our samples via GNPS [49] platform were terpenoids, indoles, carbohydrates and carbohydrate conjugates, amino acids, peptides, derivatives of purines and pyrimidines, eicosanoids and linoleic acids (Figure S3). Particularly, molecular structures of diazines, benzotraizoles, imidazopyrimidines and azides were batch-specific (Figure S3). Feature-based mass spectral molecular networking of 7,818 total MS1 molecular features (which included retention time and relative quantitative information) enabled the annotation of 248 that had matches against all publicly available reference spectra [57]. It should be noted that these are level 2 or 3 annotations according to the 2007 metabolomics standards initiative [58]. A reference-frame based approach enabled the identification of 155 features distinctly associated with specific categories relative to Ctrl group (i.e., Non-Ob/lower-dep) (Figure 3). Key molecules involved in host-microbiota interactions such as the annotation as tyrosine (level 2), a precursor of catecholamine, dopamine and serotonin, and tryptophan (level 2, cluster 14 and 26 in Figure 3), a precursor of the neurotransmitter serotonin, were depleted in Ob/higher-Dep and Ob/lower-Dep groups (Figure 2B). The amino acid, phenylalanine (Level 2, cluster 2 Figure 3), a biosynthetic precursor of tyrosine, catecholamine, dopa and dopamine was less abundant in the Ob/higher-Dep and Non-ob/higher-Dep groups, but increased with inflammation status (Figure 4A).

**Figure 3.**
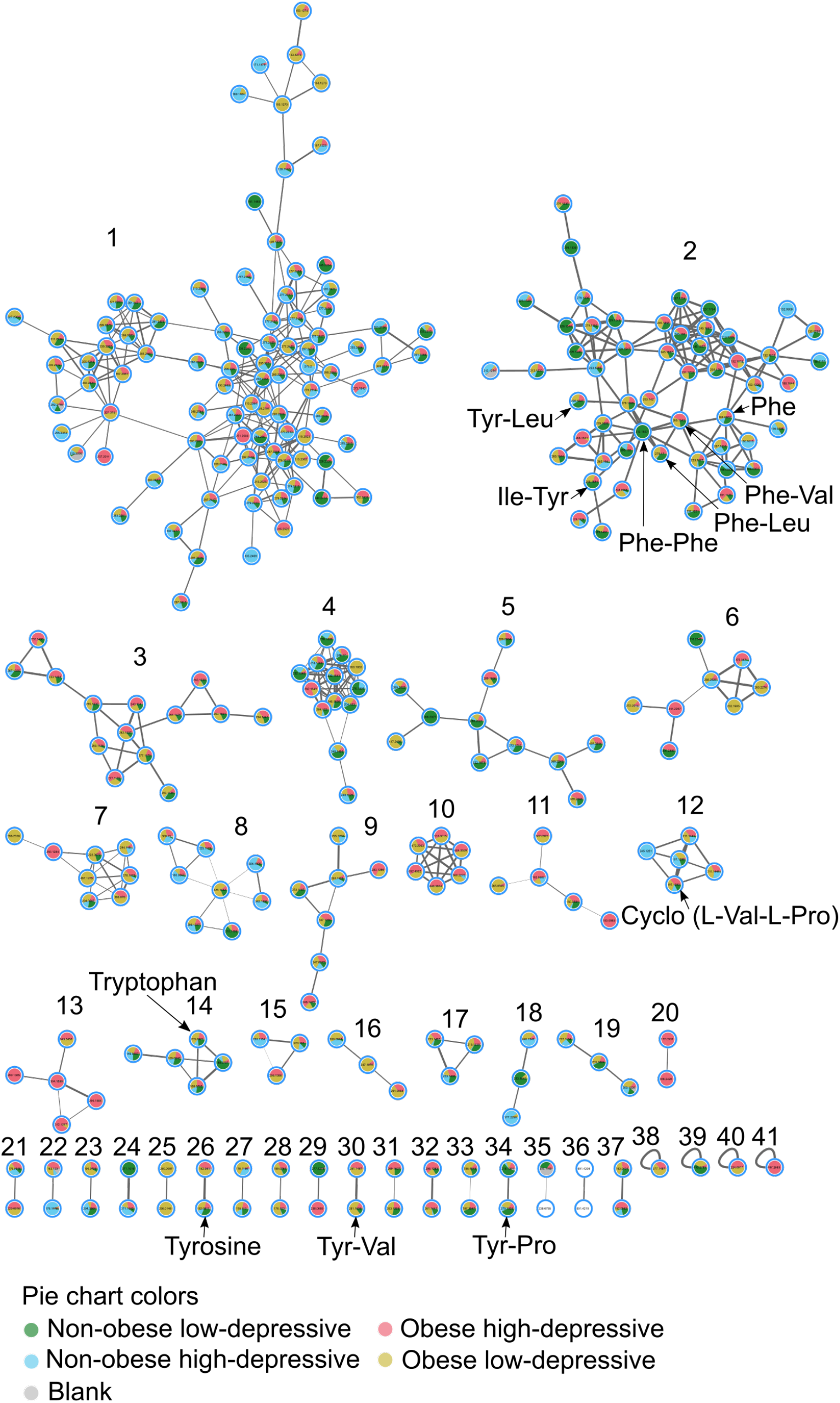
Feature-based molecular network of the ions detected in salivary metabolomes of obese-depressive group. The molecular network was generated by 293 nodes with 41 molecular clusters, which are sub-networks of a larger network generated via Global Natural Products Social Molecular Networking (GNPS). Nodes (small circles with m/z values) represent unique tandem mass spectrometry (MS/MS) consensus spectra and edges (lines) drawn between the nodes correspond to similarity (cosine score) between MS/MS fragmentation. Annotation is performed by MS/MS spectral library matching in GNPS platform. Pie charts within the individual nodes qualitatively represent specific ion presence across groups: non-obese and non- depressive, obese, depressive, and both obese and depressive symptom groups, as well as blank samples. Molecular clusters 2, 3, 4, 5, 9, 17, 19, 30 and 34 represent structural diversity of dipeptides. Molecular clusters 2, 14 and 26 represent aromatic amino acids tryptophan, tyrosine and phenylalanine.

**Figure 4.**
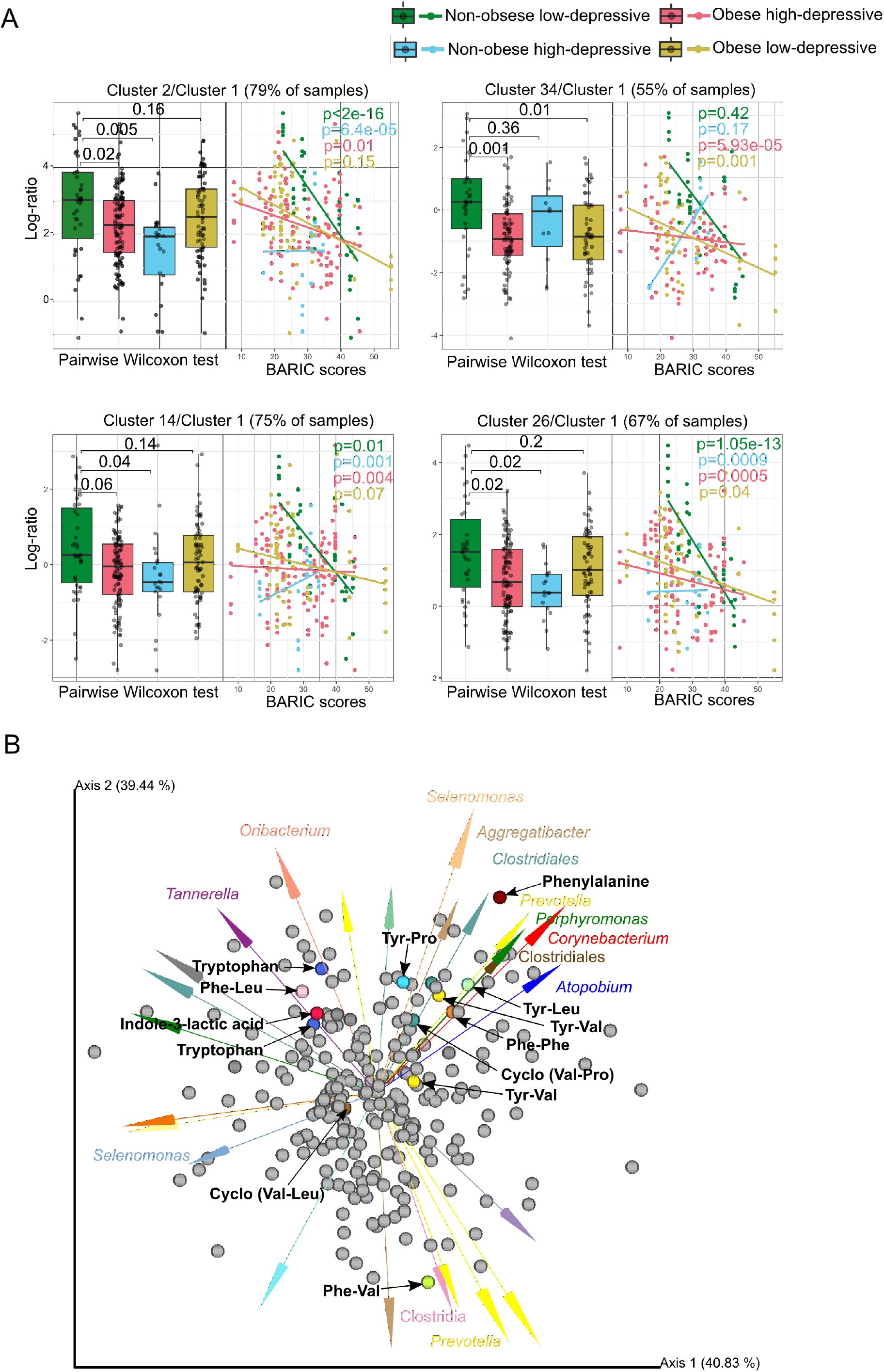
Differentially abundant molecular clusters and microbe-metabolite co-occurrences in obesity-inflammation-depressive and inflammation status. (A) Sample plot showing log-ratio of differential molecular features relative to cluster 1 (see left panel). The corresponding right panels represent a scatterplot of samples showing log-ratio of differential features versus inflammation status. Individual samples are colored by health status. Statistical significance of the log-ratios was evaluated by pairwise comparisons using Wilcoxon rank sum test. A linear regression model was employed for regressing log-ratios against BARIC values. (B) Visualization of microbe-metabolite co-occurrences. Arrows represent microbes and dots represent metabolites. The *x* and *y* axes represent principal components of the microbe-metabolite conditional probabilities as determined by the neural network. Distances between arrow tips quantify co-occurrence strengths between microbes, while directionality of the arrows indicates which microbes and metabolites have a high probability of co-occurring. Only known microbiota-derived molecules are labeled. Microbial abundances are estimated using differential abundance analysis via multinomial regression.

Within the molecular network, we also identified 41 molecular clusters primarily associated with quorum sensing molecules of microbiota, products of microbial transformation of dietary components or host molecules, and essential aromatic amino acids (Figure 3). Most intriguingly, we identified 34 structurally distinct dipeptides across groups, making it the most prevalent molecular cluster within the network (molecular features of clusters 2, 3, 5, 9, 12, 17, 19, 30, 31, 32 and 34 in Figure 3). Of these, molecular features of cluster 2 (present in 60 participants) were differentially represented in Ob/higher-Dep and Non-ob/higher-Dep individuals, while features of cluster 34 (present in 58 participants) were differentially represented in Ob/higher-Dep and Ob/lower Dep individuals, when compared to controls (see left panels in Figure 4A). Moreover, clusters 2, 14 and 26 were depleted in the Ob/higher-Dep and non-ob/higher-Dep groups, while cluster 34 was depleted in the Ob/higher-Dep and Ob/lower Dep groups. Other differentially represented molecular clusters included clusters 14 (detected in 56 participants) and 26 (detected in 58 participants), which encompassed two of the essential aromatic amino acids i.e. tryptophan and tyrosine molecules (see clusters 14 and 26 in Figure 3, Figure 4A). Molecular features from these clusters are positively associated with inflammation (right panels in Figure 4A). Abundance of features from the remaining clusters did not significantly vary across groups (data not shown). Other molecular features included previously reported microbiota-derived dipeptides (Phe-Val and Tyr-Val) (see clusters 2 and 30 in Figure 3) [42,59,60]. Dipeptide (Phe-Phe) reported to be synthesized by *Clostridium* (cluster 2 Figure 3) [61] was predominant in the Ob/higher-Dep group. Other molecules such cyclic dipeptides (Val-Pro and Val-Leu), commonly found to be made by microbes, were also identified (see cluster 2 and 12 Figure 3, Figure 4A, Table S2) [59, 60]. The majority of the other dipeptides identified were potentially related to host dietary metabolism (i.e. enzymatic digest of food proteins) [43, 44]. Among these, Tyr-Leu, Phe-Leu and Ile-Tyr (cluster 2 Figure 3), were significantly more abundant in the Ctrl group compared to the other Ob/higher-Dep and Ob/lower-Dep groups (Figure 4A) among which, Tyr-Pro (cluster 34 Figure 3) was also depleted (Figure 4A).

### Key oral microbes co-occurred with biosynthetic precursors of the neurotransmitters and dipeptide signaling molecules

Integration of the microbiome and metabolomics data revealed associations between oral microbial metabolism and key oral microbes such as *Prevotella*, Clostridia, *Selenomonas*, *Aggregatibacter*, *Oribacterium*, *Corynebacterium,* and periodontal pathogens such as *Tannerella* and *Porphyromonas* (Figure 4B). Dipeptide signaling molecules (Phe-Phe, Phe-Val and Tyr-Val) co-occurred with Clostridia, *Prevotella* and *Porphyromonas*, corroborating known associations of dipeptides produced by *Clostridium* spp. [42,59–61]. Members of Clostridia also co-occurred with phenylalanine, a potential biosynthetic precursor of dopamine, epinephrine and tryptophan. Intriguingly, *Oribacterium* belonging to *Clostridium* and *Tannerella* co-occurred with tryptophan, shown to encompass tryptophan biosynthetic pathways. Our findings further corroborate known microbial-derived cyclic dipeptides (Val-Leu and Val-Pro) associations with *Selenomonas*, *Aggregatibacter* and *Clostridium* spp. (Figure 4B) [59, 60]. Potential dietary dipeptides (Phe-Leu, Tyr-Pro and Tyr-Leu) co-occurred with *Tannerella*, *Selenomonas*, *Prevotella*, *Porphyromonas* and Clostridia [43, 44].

## Discussion

We previously reported that obesity is significantly associated with both inflammation and depressive symptoms [20,21,47]. Growing evidence also suggests that gut bacterial composition and their specialized metabolites may trigger chronic systemic inflammation in obesity- depression co-occurrences [2], highlighting the importance of the host immune and microbial interplay. In this study, we showed that the composition of salivary microbiota differ in co- occurring obesity-depressive symptoms and in relation to obesity, depression, and inflammation. We also showed that individual bacterial taxa were linked to specific host obesity-depressive symptoms ‘phenotype’, and small-molecule mediated microbe-microbe and microbe-host interactions likely play a critical role in these host phenotypes. While effects of obesity, inflammation and depression phenotypes on gut microbiome have been studied previously, this study extends our previous work [33] that identified relationships between oral microbial composition, host stress profile and inflammatory status, by providing further evidence that oral microbial composition and metabolic profiles are also influenced by the specific host phenotypes, and are likely characterized by significant alterations in the biosynthetic precursors of neurotransmitters and signaling dipeptides. These findings highlight a potential link between oral microbiota and the brain (i.e. oral-brain axis), adding to known gut microbiota-brain interactions [34–36], as well as biomarker utility of oral microbiome in studying brain and behavioral outcomes.

Examining the composition of the oral microbiome revealed significant differences based on obesity, depressive symptomatology and comorbid obesity-depressive symptomatology. At the same time, the oral microbiome composition differed by the host inflammatory processes beyond the effects of obesity or depression. This emphasizes the need of further scrutinizing the central role of microbiome-mediated inflammation in obesity-depressive symptomatology interrelationship and is closely aligned with the existing literature in chronic low-grade inflammation at the intersection of depression and obesity.

Random forest classification indicated that oral microbiota is highly predictive of obesity-depressive symptom co-occurrences, suggesting specific microbial signatures associated with obesity-depression co-occurrences. Corroborating these findings, abundances of several microbes were differentially represented across the obesity-depressive symptomatology groups as revealed by the differential abundance analysis. Gram-negative microbes have been shown to be associated with inflammation due to their LPS cell wall, the hallmark trait of Gram-negative bacteria. We found that Gram-negative microbes *Prevotella*, *Aggregatibacter*, *Pseudomonas*, *Campylobacter*, *Selenomonas*, *Leptotrichia*, *Capnocytophaga*, and Gram-negative periodontal pathogens such as *Treponema*, *Veillonella*, *Porphyromonas* and *Fusobacterium* are enriched in Ob/higher-dep group. However, we found no significant correlation with BARIC scores that measured monocytes’ responsiveness to a β-AR agonist during an inflammatory response to LPS, indicating inflammation regulatory processes [47]. Increased abundance of *Prevotella* in the human oral cavity has been previously ambiguously associated with both health and disease conditions [26,62,63]. Pathogenic *Campylobacter* has been shown to increase anxiety-like behavior in mice [64] and *Aggregatibacter* has been reported to be associated with inflammation. Notably, Gram-positive beneficial microbes *Bifidobacterium* and *Lactobacillus* depleted in Ob/higher-Dep group are in line with their activity as they are reported to exhibit antidepressant and anti-obesity effects, and reduced levels of TNF-α in both clinical and animal studies [65–67]. All of these differentially abundant oral taxa present potential biomarkers in obesity-depression co-occurrences, however, more studies are needed to further confirm these findings, as our study did not find significant differences in the abundances of microbes at genera-level.

We also found differences in relative abundance patterns in many molecules across the obesity-depression symptoms groups, including quorum sensing molecules of microbiota, products of microbial transformation of dietary components or host molecules and aromatic amino acids. Importantly, metabolites of aromatic amino acids tryptophan and tyrosine, both of which are precursors of the neurotransmitter serotonin, have been mechanistically implicated in obesity-depression associations [68], and play signaling roles in host-microbe interactions in the gut [69], were depleted in obese individuals compared to the control group. Host dietary dipeptides (Tyr-Leu and Phe-Leu) that were significantly less abundant among the obese individuals compared to the control group in this study are shown to display anti-depressant-like activity as greater abundance of Tyr-Leu activates serotonin, dopamine and gamma aminobutyric acid (GABA) receptors in mice [43, 44]. Tyr-Pro and Ile-Tyr, which were also depleted in the obese individuals in our study, are an inhibitor of angiotensin I-converting enzyme (ACE) with antihypertensive activity [70] and affect catecholamine (e.g. dopamine and noradrenaline) metabolism in the mouse brain [71], respectively. These findings offer initial mechanistic insight into comorbid obesity and depression, albeit complex.

Furthermore, we identified several structurally distinct dipeptides that were positively associated with inflammation. To our knowledge, it is the first time that microbial-derived dipeptide (Phe-Val, Tyr-Val and Phe-Phe) and cyclic dipeptides signaling molecules (Val-Pro and Val-Leu) were detected in salivary metabolomes. Biosynthetic gene clusters and the production of dipeptides (Phe-Val and Tyr-Val) have been recently identified in the human microbiome [42,59,60]. These molecules are known to play key roles in quorum sensing (cell-to- cell communication to maintain cell density) and virulence, and promote growth of beneficial *Bifidobacterium* [41]. A previous study showed that Phe-Phe derived from *Clostridium sp.* can inhibit host proteins by chemical modification of the host cellular proteins, especially by targeting cathepsins in human cell proteomes [61]. Given our findings that Phe-Phe was highly abundant in the Ob/higher-Dep group, its biological role in the cellular inflammatory process which likely underlie obesity-depression comorbidity warrants further investigation.

Our findings of specific microbe-metabolite interactions with potential to influence host’s brain functioning offer potentially significant insight into the role of host immune-microbiome interplay in comorbid obesity-depression and is likely through microbial neurotransmitters. Metabolic pathways for biosynthesis of neuroactive molecules in the genomes of human- associated genera *Clostridium* and *Tannerella* have been recently reported [35]. Intriguingly, members of *Clostridium* and *Tannerella* co-occurred with tryptophan and have been detected/reported to harbor genes for tryptophan biosynthesis [35]. Members of Clostridia co-occurred with phenylalanine, a potential biosynthetic precursor of dopamine, epinephrine and tryptophan, have been shown to be key species in neuropsychiatric disorders and shown to produce dopamine in mice [36, 72]. Many of these molecules including the dipeptides, shown to have potential to cross the intestinal barrier and blood brain barrier, may modulate the oral–brain connection through neurotransmitter signaling pathways [35, 72]. Such neurotransmitters and their biosynthetic precursors may offer promising targets for therapeutics.

There is a caveat in this study that merits caution: in an effort to recruit individuals with subclinical levels of depressive mood co-occurring with a range of obesity without antidepressant intake or heterogeneous clinical depression, the participants exhibited low levels of BDI scores on average which may limit the applicability of our findings to clinical depression. At the same time, it is notable that host-microbiome-metabolome signatures and their interactions appear to be salient in pathophysiology of subclinical depression symptomatology. We also acknowledge a small sample size of the study participants, in spite of the expanded specimen sample size owing to multiple saliva collections.

## Conclusions

Despite these limitations, our study significantly expands the evidence for microbial specialized metabolites and peptides with neuroactive potential, adding further research avenues into microbiome-host physiology interactions and there is a great deal of clinical potential in understanding and modifying these interactions. Furthermore, it provides initial evidence for a foundation of the microbial oral-brain axis in addition to the gut-brain axis in the context of obesity-depression-inflammation interrelationships.

## Data Availability

Sample metadata, the raw and processed 16S sequencing data and their associated feature tables, and preparation metadata are available in Qiita Study ID 11259 (https://qiita.ucsd.edu/study/description/11259). Mass spectral files and LC-MS/MS preparation metadata are accessible from the MassIVE repository accession ID MSV000083077 (ftp://massive.ucsd.edu/MSV000083077). The GNPS feature based molecular networking job is available at https://gnps.ucsd.edu/ProteoSAFe/status.jsp?task=f192a0030f694224a0ba8f08223a1323

ftp://massive.ucsd.edu/MSV000083077

https://gnps.ucsd.edu/ProteoSAFe/status.jsp?task=f192a0030f694224a0ba8f08223a1323

## Declarations

### Ethics approval and consent to participate

All participants provided informed consent to the protocol prior to the commencement of the study. The Ethics Committee of the University of California, San Diego, CA, USA, approved the study design as well as the procedure for obtaining informed consent (IRB reference number: 171027). All experiments were performed in accordance with the approved guidelines of UCSD Human Research Protections Program.

### Consent for publication

Not applicable

### Competing interests

PCD serves as a scientific advisor to Sirenas, Cybele and Galileo. PCD is also a founder and scientific advisor of Ometa and Enveda with approval by UC San Diego.

### Funding

This work was supported in part by R01 HL90975 and 90975S1 from the NIH (Hong), a Seed Grant from the Center for Microbiome Innovation at UC San Diego (Hong), Wayne State University Endowment Fund (Hong), and the Kavli Institute for Brain and Mind (KIBM) Innovative Research Grant (Aleti).

### Authors’ contributions

Hong designed and obtained funding for the study. GA performed the data analysis. JNK, KW, AT, ADS and Huang assisted with the data analysis. GA, Hong, PCD, ADS and RK interpreted the results. GA, ET and Hong wrote the original manuscript. GA, JNK, ET, KW, AT, Huang, ADS, Hong, PCD and RK reviewed and edited the manuscript.

## Acknowledgements

Not applicable

## Supplementary Materials and Methods

### Blood collection and cellular inflammation assay

Blood samples were obtained for all participants after 12h of fasting except for plain water and collected in heparin anti-coagulant vacutainers (BD, Franklin Lakes, NJ). Cellular inflammation regulation assays were performed on heparinized whole blood within 1h of collection. Briefly, 200 pg/mL of lipopolysaccharide (LPS) (E.coli 0111:B4, catalog #L4391, Sigma-Aldrich, St. Louis, MO) was added to 300 μL of blood in sterile 96-well polypropylene cell culture plates andL of blood in sterile 96-well polypropylene cell culture plates and incubated for 30 min at 37°C with 5% CO2. Media-treated samples served as controls. This exogenous LPS dose was previously determined to elicit significant activation of monocytes, with 30-90% producing TNF-α [1]. Monocyte beta-adrenergic receptor-mediated inflammation control (i.e., “BARIC”) was determined based on the inhibitory effect of isoproterenol (Iso), a non-specific β1/2AR agonist, on monocytic intracellular TNF-α production in LPS-stimulated blood as aforementioned. Briefly, LPS-stimulated blood was incubated with isoproterenol in 10^-8^ M final concentration and evaluated for intracellular monocyte TNF-α production using flow cytometry, as previously described [1]. The proportion of CD14^+/dim^HLA-DR^+^ (CD14: cat. #301808; HLA-DR: cat. #307606, BioLegend, San Diego, CA) cells that were TNF-α^+^ was determined using FlowJo software (v10, TreeStar, Ashland, OR), and gates adjusted for TNF-α- stained sample via fluorescence-minus-one controls [2, 3]. Ultimately, BARIC was calculated as the arithmetic difference in %TNF-α+ monocytes between LPS-treated and LPS+isoproterenol-treated samples. Greater BARIC values indicate greater β-AR responsivity, and thus, better Iso/β-AR-mediated inflammation regulation. Smaller BARIC values may indicate impairment in cellular pathways that regulate inflammatory responses mediated by β-ARs (e.g., diminished receptor sensitivity to agonists). BARIC measures monocytes responsivity to a β-AR agonist during an inflammatory response to LPS. Reduced BARIC has been associated with hypertension, cardiovascular disease risk factors, obesity, and higher serum cytokine levels [2, 3].

### Saliva collection, DNA extraction and 16S sequencing

Saliva collection procedure and 16S sequencing data was published previously [2]. However, obesity-depressive symptom relationships were not previously investigated, and instead had focused on temporal variation of the oral microbiota. Briefly, participants were provided with Salivette (Sarstedt, #51.1534, Nümbrecht, Germany) to roll the cotton Salivette inside the mouth to stimulate salivation without chewing. Saturated Salivette was placed back into the tube by mouth. Salivettes from each participant were collected at five time points across a single day: waking, mid-morning (10:00 hrs), midday (12:00 hrs), afternoon (14:00 hrs), and evening (17:00 hr). All waking samples were collected prior to oral hygiene activity, and ingestion of food or drink. In addition, participants were instructed to abstain from consuming food or drinks other than plain water for 30 min and to rinse their mouth with water prior to collection at all other time points. Next, saliva was recovered from Salivette tubes by centrifuging at 1,000 x g for 2 minutes at 4°C and stored at -80°C. DNA from saliva samples was extracted by employing Qiagen PowerSoil DNA kit as previously described [4]. V4 region of the 16S gene was amplified according to the Earth Microbiome Project protocol [5, 6] and sequenced on the Illumina MiSeq sequencing platform with a MiSeq Reagent Kit v2 and paired-end 150 bp cycles.

### 16S sequencing data processing

Sequences were demultiplexed based on the barcode associated with each sample and sequence quality control and ASV (Amplicon Sequence Variants) feature table construction was conducted using the Deblur algorithm in QIIME2 (v.2018.4) [7]. Next, 223 potential sequencing contaminants that appeared in both true and blank samples were removed from the ASV table using *decontam* in R [8]. Low abundance features with fewer than 10 reads across samples and singleton features present only in one sample were excluded. Taxonomy assignment was performed by employing QIIME2 feature-classifier plugin with a pre-fit classifier [9] for the 99% reference tree of Greengenes 13_8 database. The output feature table contained an average of 19,412 ± 9,187 sequences per sample after removal of mitochondrial and chloroplast-derived sequences. Multiple rarefactions were computed to a minimum depth of 1,122 reads to mitigate uneven sequencing depth across samples. This resulted in 257 samples with 1,516 unique features/ASVs and 455 unique taxa. Next, alpha-diversity indices Shannon diversity index and Faith’s Phylogenetic Diversity were calculated. Beta-diversity, was calculated using unweighted UniFrac distance, which reflects presence-absence of taxa. We performed ordination on output distance matrices using principal coordinates analysis (PCoA) and following visualization using EMPeror plugin in QIIME2 [10].

### Small molecule metabolites detection through mass spectrometry

Saliva was dried and resuspended in 80% MeOH−20% water (Optima LC-MS grade; Fisher Scientific, Fair Lawn, NJ, USA). Untargeted metabolomics was conducted with an ultrahigh- performance liquid chromatography (Vanquish; Thermo Fisher Scientific, Waltham, MA, USA) system coupled to an orbitrap mass spectrometer (QExactive, Thermo Fisher Scientific). A C18 reversed-phase UHPLC column (Kinetex, 1.7-µm particles size, 50 x 2.1 mm) (Phenomenex, Torrance, CA, USA) was used for chromatographic separation. A linear gradient was applied as follows: 0 to 0.5 min, isocratic at 5% mobile phase (MP) B; 0.5 to 8.5 min, 100% MP B; 8.5 to 11 min, isocratic at 100% MP B; 11 to 11.5 min, 5% MP B; 11.5 to 12 min, 5% MP B, where mobile phase A is water with 0.1% formic acid (vol/vol) and mobile phase B is acetonitrile−0.1% formic acid (vol/vol) (LC-MS grade solvents; Fisher Chemical). Electrospray ionization in the positive mode was used. MS spectra were acquired in the mass range of m/z 100 to 2,000.

### MS1 feature finding and data processing

Raw QExactive files were converted to .mzXML format using ProteoWizard tool MSConvert [11] software. Data quality was assessed by evaluating the m/z error and retention time of the LC-MS standard solution (i.e., mixture of six compounds). MS1 feature finding was performed in MZmine2 preprocessing workflow (MZmine-2.37.corr17.7_kai_merge2 version) available at (https://github.com/robinschmid/mzmine2/releases) [12]. The mzMINE parameters used for feature finding are as follows: mass detection (centroid; MS1, 1.5E3; MS2, 90); ADAP Chromatogram builder (minimum group size in number of scans, 4; group intensity threshold, 5E3; minimum highest intensity, 2E3; m/z tolerance, 0.001 m/z to 20 ppm); chromatogram deconvolution (local minimum search, chromatographic threshold of 96%, search minimum in retention time [RT] range [minutes] of 0.03, minimum relative height of 5%, minimum absolute height of 2E3, minimum ratio of peak top/edge of 1 and peak duration range [minutes] of 0 to 2;m/z center calculation set to auto; m/z range for MS2 scan pairing (daltons) of 0.02 and RT range for MS2 scan pairing (minutes) of 0.15); isotope peaks grouper (m/z tolerance set to 0.0015 m/z or 10 ppm; retention time tolerance of 0.05, maximum charge of 3; and representative isotope set to most intense); order peak lists; join aligner (m/z tolerance set at 0.0015 m/z or 15 ppm; weight for m/z of 2; retention time tolerance of 0.2 min; weight for RT of 1. A filter was used such that only features present in at least two samples were included.

### Feature based mass spectral molecular networking (FBMN)

The output of aforementioned workflow, a data matrix of MS1 features that triggered MS2 scans by sample (.mgf and .csv quant table), were uploaded along with the metadata file to Global Natural Product Social Molecular Networking (GNPS) (https://gnps.ucsd.edu) [13, 14]. Feature- based molecular networking (version release_20) [15] was performed, and library IDs were generated. Molecular networking parameters were set as follows: precursor ion mass tolerance and fragment ion tolerance of 0.02 Da to cluster consensus spectra; the minimum score between a pair of MS2 consensus spectra was set at 0.7 and 6 as the minimum number of ions matched as described at https://gnps.ucsd.edu/ProteoSAFe/status.jsp?task=f192a0030f694224a0ba8f08223a1323. The molecular network output from GNPS was then uploaded to Cytoscape (version 3.5.1 http://www.cytoscape.org/) [16], for advanced visualization. Nodes were labelled with spectral matches to GNPS with m/z values, and edge thickness is proportional to the cosine score.

## Supplementary figures

**Figure S1.**
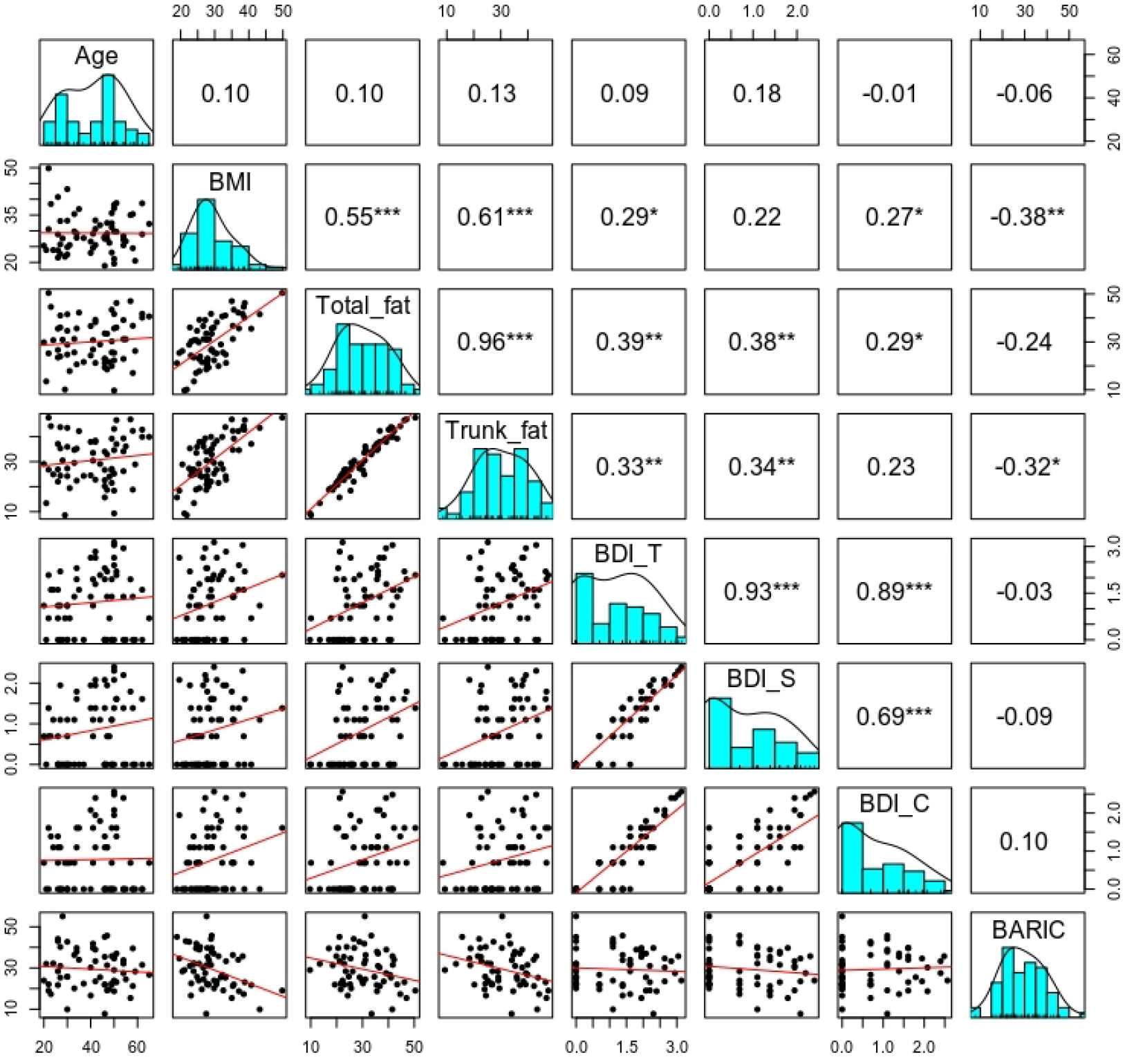
Matrix of plots illustrating Pearson correlations among obesity, depressive symptoms, inflammation and sex, across participants. Histograms of the variables displayed along the matrix diagonal represent distribution of samples and scatter plots of variable pairs are displayed in the off diagonal. Correlation coefficients displayed represent the slopes of the least- squares reference lines in the scatter plots.

**Figure S2.**
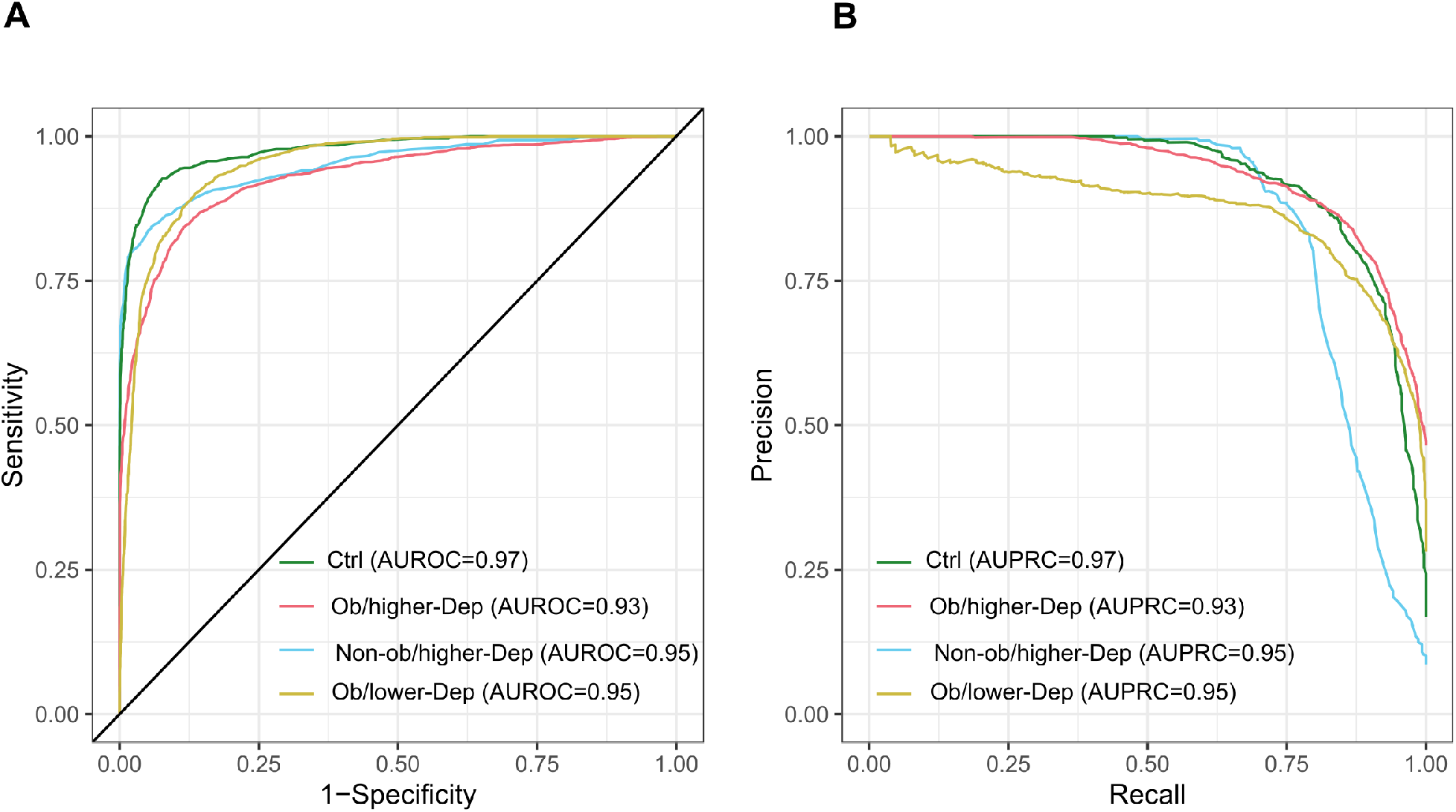
Per sample based RF analysis. (A), Receiver operating characteristic curves (AUROC) illustrating classification accuracy of the random forest model across all groups (i.e. controls, Ob/lower Dep, Non-ob/higher-Dep, Ob/higher-Dep) and (B), Area under precision recall curves (AUPRC) illustrating performance of the random forest model across all groups.

**Figure S3.**
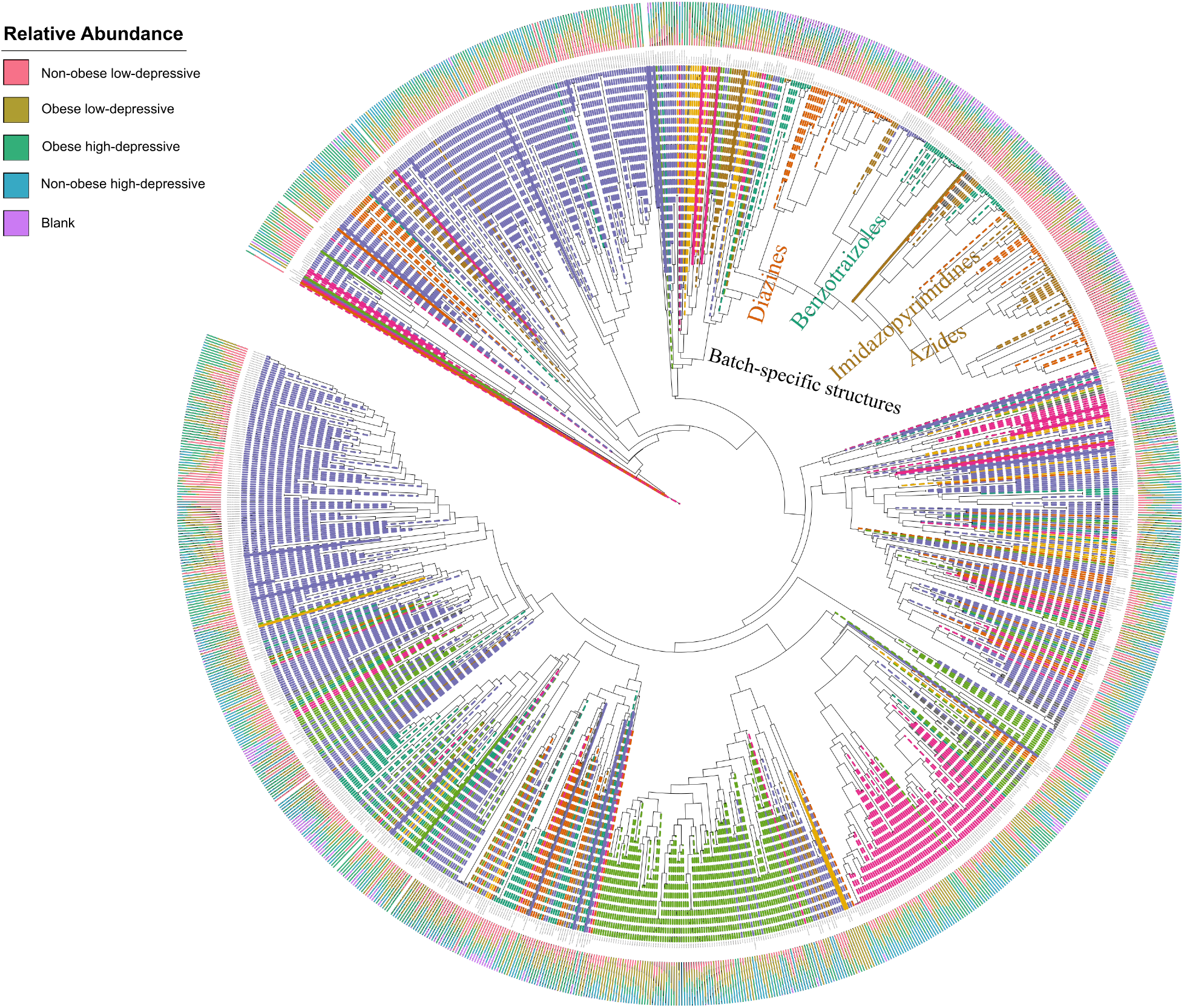
Chemical diversity captured in salivary metabolomes. Branches in the circular chemical tree are colored according to the class type and branch labels represent putatively annotated chemical features at subclass level based on chemical taxonomy. Bar graphs at the leaf tips illustrate relative abundance of molecules across groups.

