## Supplementary methods and figures for "Salivary bacterial signatures in depression-obesity comorbidity are associated with neurotransmitters and neuroactive dipeptides"

### Supplementary figures

**Figure S1**. Matrix of plots illustrating Pearson correlations among obesity, depressive symptoms, inflammation and sex, across participants. Histograms of the variables displayed along the matrix diagonal represent distribution of samples and scatter plots of variable pairs are displayed in the off diagonal. Correlation coefficients displayed represent the slopes of the least-squares reference lines in the scatter plots.

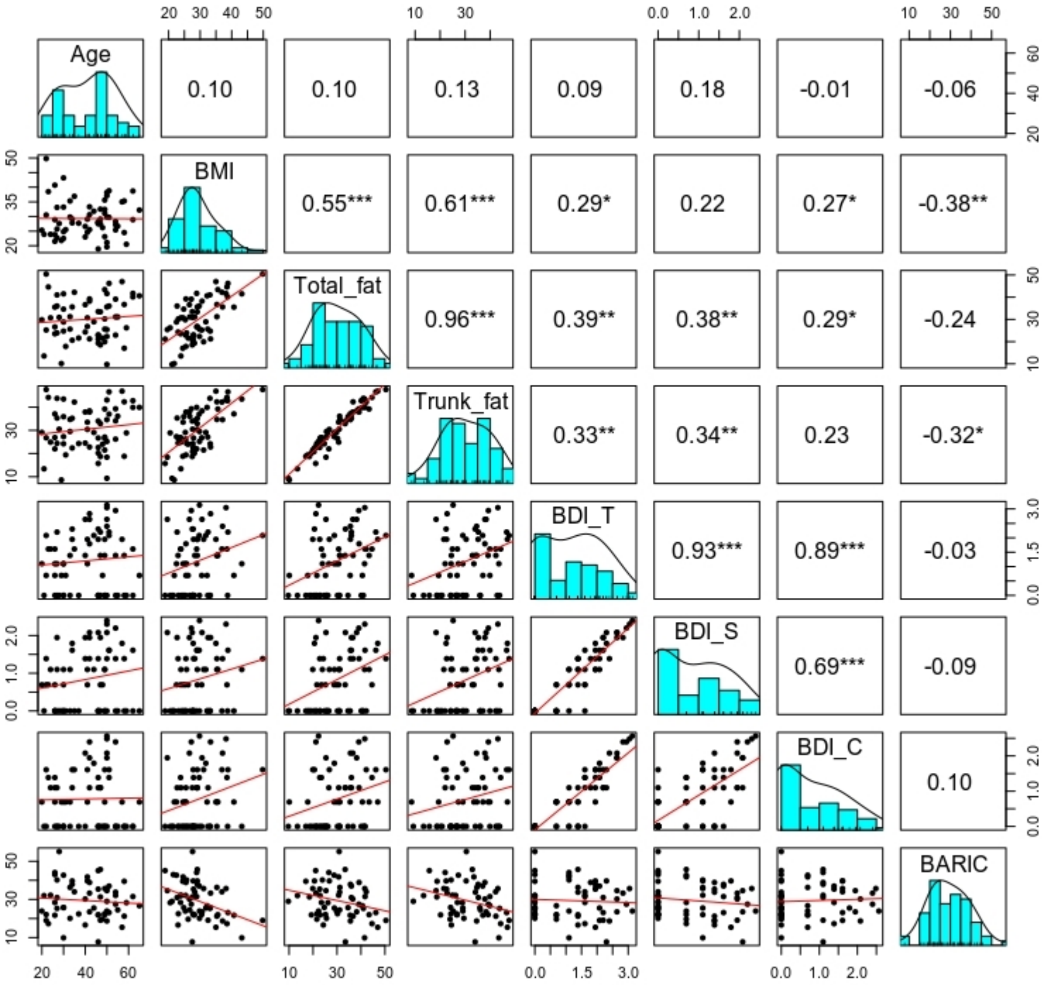

**Figure S2**. Per sample based RF analysis. (A), Receiver operating characteristic curves (AUROC) illustrating classification accuracy of the random forest model across all groups (i.e. controls, Ob/lower Dep, Non-ob/higher-Dep, Ob/higher-Dep) and (B), Area under precision recall curves (AUPRC) illustrating performance of the random forest model across all groups.

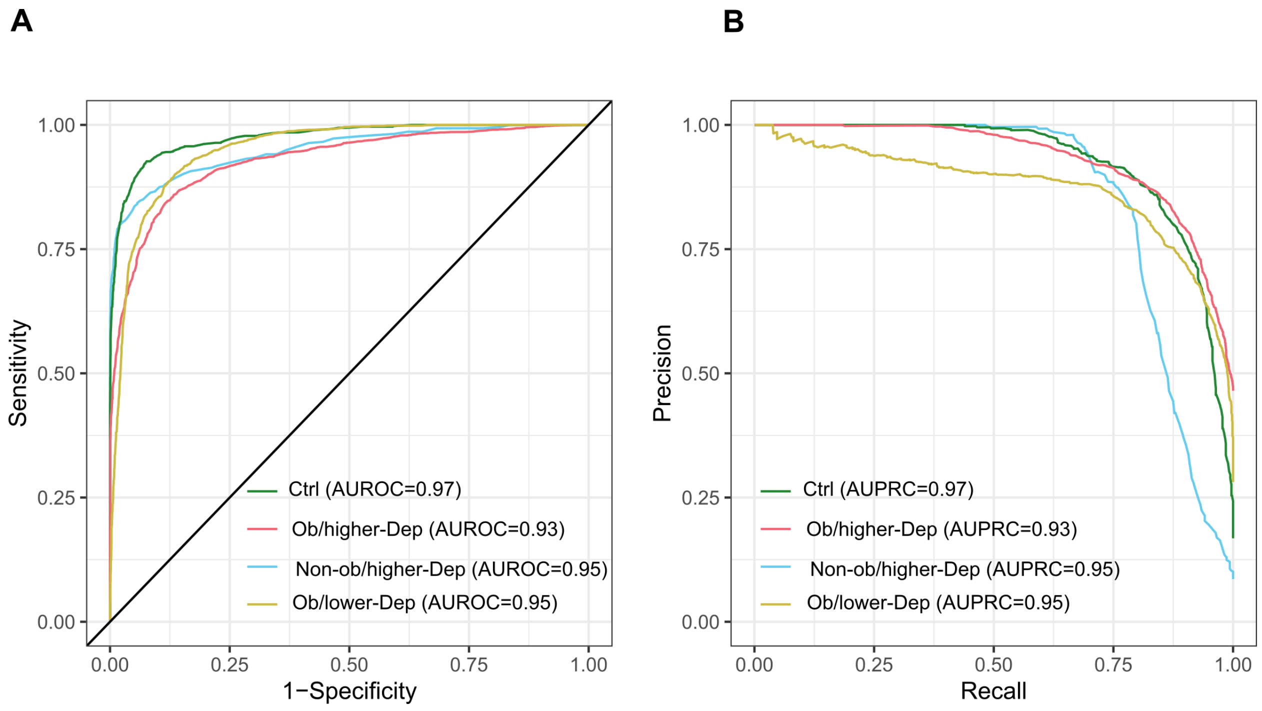

**Figure S3**. Chemical diversity captured in salivary metabolomes. Branches in the circular chemical tree are colored according to the class type and branch labels represent putatively annotated chemical features at subclass level based on chemical taxonomy. Bar graphs at the leaf tips illustrate relative abundance of molecules across groups.

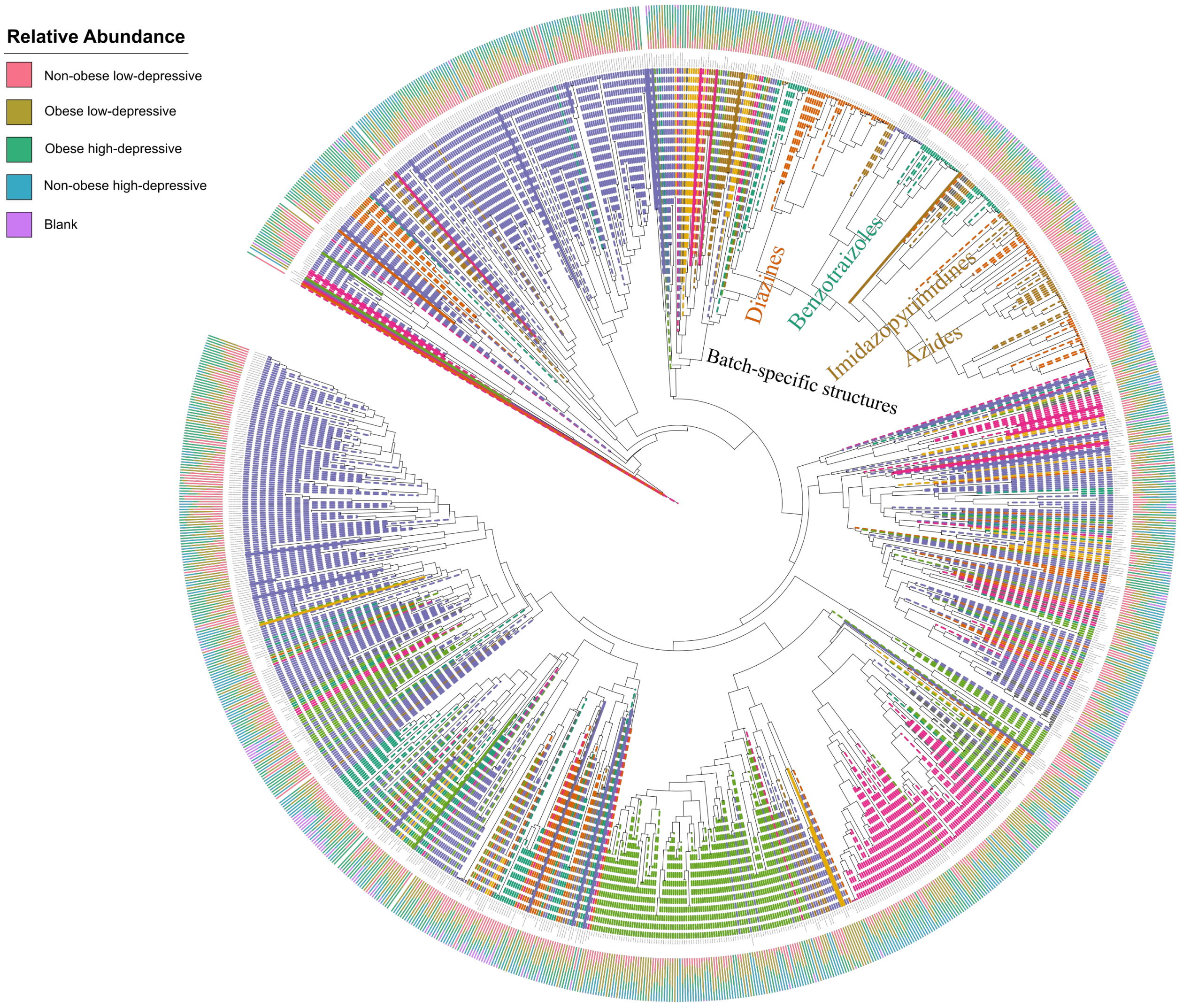

11. Chambers MC, MacLean B, Burke R, Amodei D, Ruderman DL, Neumann S, et al. A cross-

platform toolkit for mass spectrometry and proteomics. Nat. Biotechnol. Nat Biotechnol; 2012. p. 918–20.

12. Pluskal T, Castillo S, Villar-Briones A, Orešič M. MZmine 2: Modular framework for processing, visualizing, and analyzing mass spectrometry-based molecular profile data. BMC Bioinformatics. BMC Bioinformatics; 2010;11.

13. Aron AT, Gentry EC, McPhail KL, Nothias LF, Nothias-Esposito M, Bouslimani A, et al. Reproducible molecular networking of untargeted mass spectrometry data using GNPS. Nat Protoc. Nature Research; 2020;15:1954–91.

14. Wang M, Carver JJ, Phelan V V., Sanchez LM, Garg N, Peng Y, et al. Sharing and community curation of mass spectrometry data with Global Natural Products Social Molecular Networking. Nat. Biotechnol. Nature Publishing Group; 2016. p. 828–37.

15. Nothias LF, Petras D, Schmid R, Dührkop K, Rainer J, Sarvepalli A, et al. Feature-based molecular networking in the GNPS analysis environment. Nat Methods. Nature Research; 2020;17:905–8.

16. Shannon P, Markiel A, Ozier O, Baliga NS, Wang JT, Ramage D, et al. Cytoscape: A software Environment for integrated models of biomolecular interaction networks. Genome Res. Genome Res; 2003;13:2498–504.
